# A human multisystem disorder with autoinflammation, leukoencephalopathy and hepatopathy is caused by mutations in *C2orf69*

**DOI:** 10.1101/2021.03.08.21252805

**Authors:** Eva Lausberg, Sebastian Gießelmann, Joseph P Dewulf, Elsa Wiame, Anja Holz, Ramona Salvarinova, Clara Van Karnebeek, Patricia Klemm, Kim Ohl, Michael Mull, Till Braunschweig, Joachim Weis, Clemens Sommer, Stephanie Demuth, Claudia Haase, François-Guillaume Debray, Cecile Libioulle, Daniela Choukair, Prasad T. Oommen, Arndt Borkhardt, Harald Surowy, Dagmar Wieczorek, Robert Meyer, Thomas Eggermann, Matthias Begemann, Emile Van Schaftingen, Martin Häusler, Klaus Tenbrock, Lambert van den Heuvel, Miriam Elbracht, Ingo Kurth, Florian Kraft

## Abstract

**Background:** Deciphering the function of the many genes previously classified as uncharacterized “open reading frame” (orf) completes our understanding of cell function and its pathophysiology.

**Methods:** Whole-exome sequencing, yeast 2-hybrid and transcriptome analyses together with molecular characterization are used here to uncover the function of the *C2orf69* gene.

**Results:** We identify loss-of-function mutations in the uncharacterized *C2orf69* gene in eight individuals with brain abnormalities involving hypomyelination and microcephaly, liver dysfunction and recurrent autoinflammation. C2orf69 contains an N-terminal signal peptide that is required and sufficient for mitochondrial localization. Consistent with mitochondrial dysfunction, patients show signs of respiratory chain defect and a CRISPR-Cas9 knockout cell model of *C2orf69* shows comparable respiratory chain defects. Patient-derived cells reveal alterations in immunological signaling pathways. Deposits of PAS-positive material in tissues from affected individuals together with decreased glycogen branching enzyme 1 (GBE1) activity indicate an additional impact of C2orf69 on glycogen metabolism.

**Conclusion:** Our study identifies C2orf69 as an important regulator of human mitochondrial function and suggests an additional influence on other metabolic pathways.

**Graphical Abstract:** 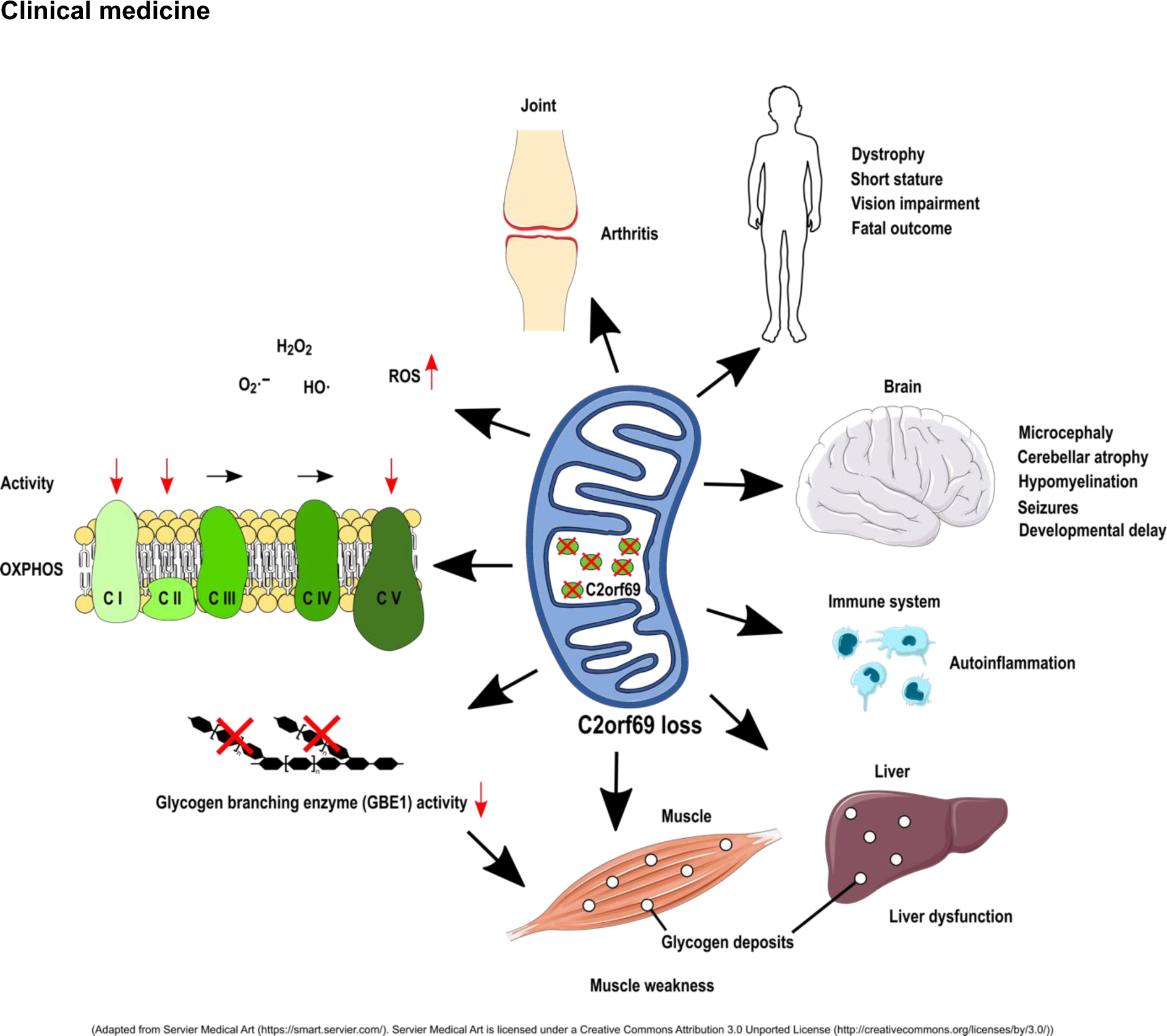

## Introduction

Inborn errors of metabolism (IEM) are a genetically heterogeneous group of more than 1000 diseases (1). They result from metabolic defects due to deficiency of enzymes, membrane transporters, or other functional proteins. IEMs can fall into many categories with defects in carbohydrate or protein metabolism, fatty acid oxidation, glycogen storage, or dysfunction of peroxisomes, lysosomes, or mitochondria (2). Initially considered rare, they have a significant global birth prevalence of approximately 1 in 2,000 (3). Due to the large number of clinically overlapping disorders and manifold manifestations, IEMs are often difficult to diagnose. Specific signs can be absent and clinical symptoms range from mild developmental delay to severe metabolic acidosis with periods of remission or sudden death. A large class of IEMs directly affects the energy metabolism, especially the function of mitochondria which are crucial for ATP supply via oxidative phosphorylation (OXPHOS) (4). The mitochondrial proteome consists of about 1500 proteins, many of which are still uncharacterized, leaving an incomplete understanding of mitochondrial physiology (5). We here identify homozygous loss-of-function mutations in the uncharacterized *C2orf69* gene as a cause for a complex multisystem disease and describe an important role for this gene in mitochondrial metabolism.

## Results

### Mutations in the uncharacterized C2orf69 gene cause a multisystem human disease

We investigated a male index patient (patient I), born to consanguineous parents who suffered from severe developmental delay, hepatopathy, recurrent septic and aseptic inflammations of joints starting early after birth and followed by an aseptic pericarditis at around one year of age. The episodes of inflammation were accompanied by elevated C-reactive protein (CRP) and partially responsive to glucocorticoids. The severe dystrophy made feeding via percutaneous gastrostomy necessary. Brain MRI revealed a prominent cerebellar atrophy, thin corpus callosum and diffuse hypomyelination (**Supplement Figure 1**). He showed muscle weakness, general dystrophy and a short stature, thyroid dysfunction, biventricular cardiac hypertrophy and osteopenia. A severe epilepsy finally made ACTH therapy necessary, which also reduced the autoinflammation. With about 2-3 years he died from multi-organ failure (**Supplement Table 1**). Whole-exome sequencing (WES) revealed a potentially disease-causing homozygous 1-bp deletion (NM_153689.6:c.298del) in *chromosome 2 open reading frame 69* (*C2orf69*), a hitherto uncharacterized gene (**Figure 1A**). The variant results in a premature stop codon (p.(Gln100Ser*fs*\*18)). Both healthy parents and the two unaffected siblings were heterozygous for the *C2orf69* deletion. *C2orf69* consists of two exons and encodes a protein that is highly conserved among vertebrates and *Drosophila*. According to *in silico* predictions the gene product has an N-terminal 24 amino acid signal peptide (SP) and a protein-family domain of unknown function (UPF0565) with phosphorylation-, ubiquitination-, SUMOylation- and NEDDylation-sites (6-9) (**Figure 1B**).

**Figure 1.**
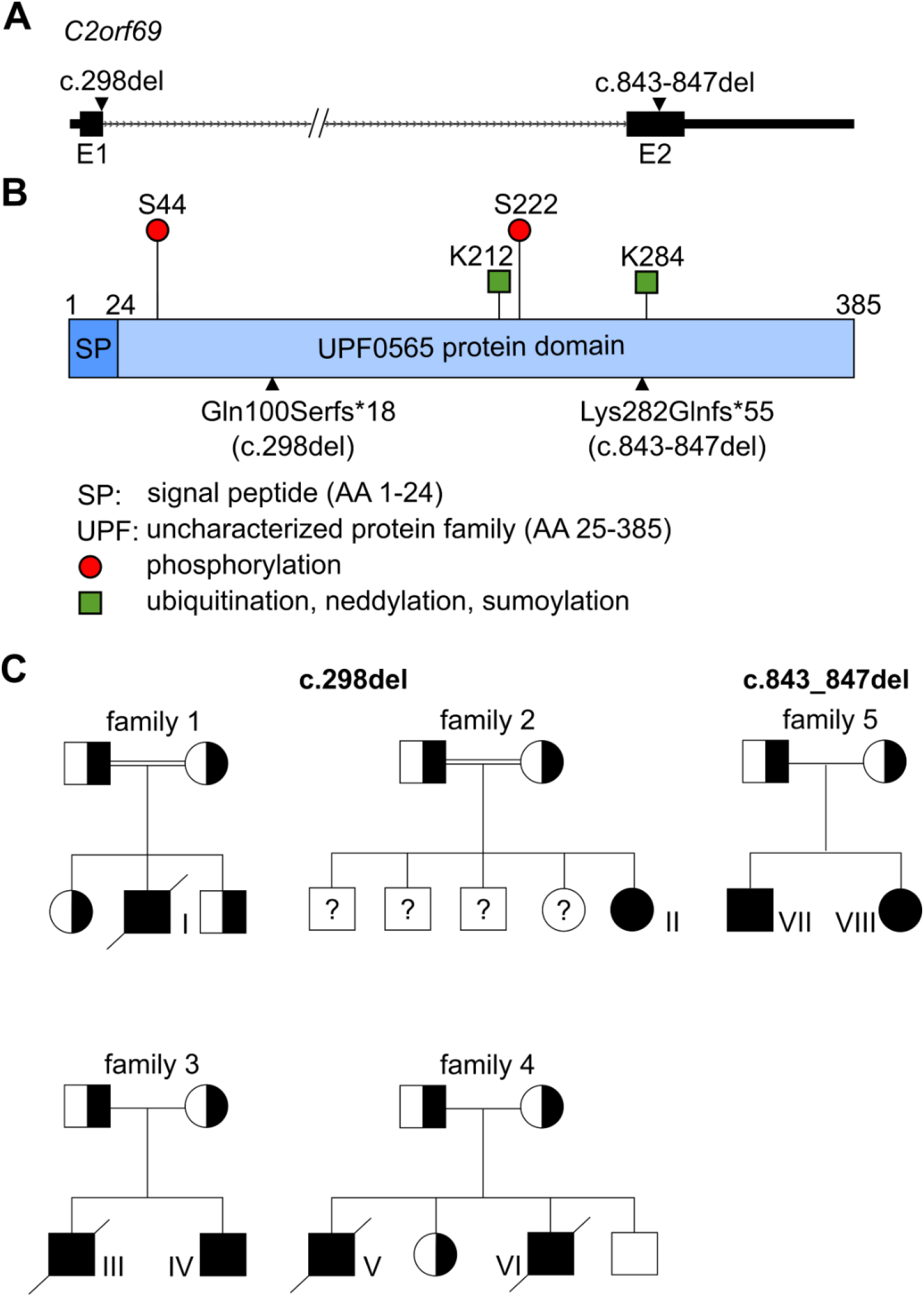
Gene and protein structure of C2orf69. (A) *C2orf69* is composed of two exons (boxes E1 and E2). 3’- and 5’-UTR are indicated as thick line, intron region as arrowed line. Triangles present the mutations found in the patients. (B) Domain architecture of the human C2orf69 protein. The protein harbors a 24 amino acid (AA) signal peptide (SP) and an uncharacterized UPF0565 protein domain. Circles and squares indicate post-transcriptional modifications and triangles the mutation sites. (C) Pedigrees of families show the segregation of the C2orf69 variants. The homozygously mutated allele is shown in black, the wild type in white and heterozygous carriers as split symbol.

Through GeneMatcher (10) and existing collaborations we identified seven additional patients from four independent families with homozygous loss-of-function mutations in *C2orf69* (**Figure 1C**). Five patients from three different families harbored the same homozygous c.298del mutation in *C2orf69*, whereas two patients from another family showed a homozygous 5-bp deletion (c.843_847del), resulting in a premature stop codon (p.(Lys282Gln*fs*\*55)). Both variants are not listed in public databases and no homozygous loss-of-function variants of C2orf69 are known so far. All eight individuals showed very similar clinical signs and brain MRI findings were remarkably consistent (**Supplement Table 1, Supplement Figure 1**). Different degrees of frontotemporal atrophy, hypomyelination, thin corpus callosum and Dandy Walker variant with hypoplasia of the caudal vermis were typical findings in MRI. Patients I, III, V and VI died at a young age. Interestingly, four patients (patient III-VI) were initially suspected to have a glycogen storage disease (GSD), because glycogen branching enzyme (GBE1) activity was absent or reduced in tissues (**Supplement Table 2**). In accordance, PAS-positive deposits were observed in myocytes and macrophages of patient III, and liver of patients III, V and VI. The autopsy of patient I showed PAS-positive accumulations in hepatocytes, cardiomyocytes and macrophages (**Figure 2A-F, Supplement Figure 1D**). These deposits were resistant to digestion with amylase (PAS diastase stain) (**Figure 2B-C, Figure 2E-F**), which led to the assumption of GSD type IV (GSD IV), although no pathogenic variants in the *GBE1* gene were present. The muscle biopsy of patient III showed polyglucosan bodies in the electron microscopy (EM) (**Figure 2G-H)**. Disintegration of several mitochondria accompanied by osmiophilic membranous material in the mitochondria indicated disturbed mitophagy in the muscle (**Figure 2I-J**).

**Figure 2.**
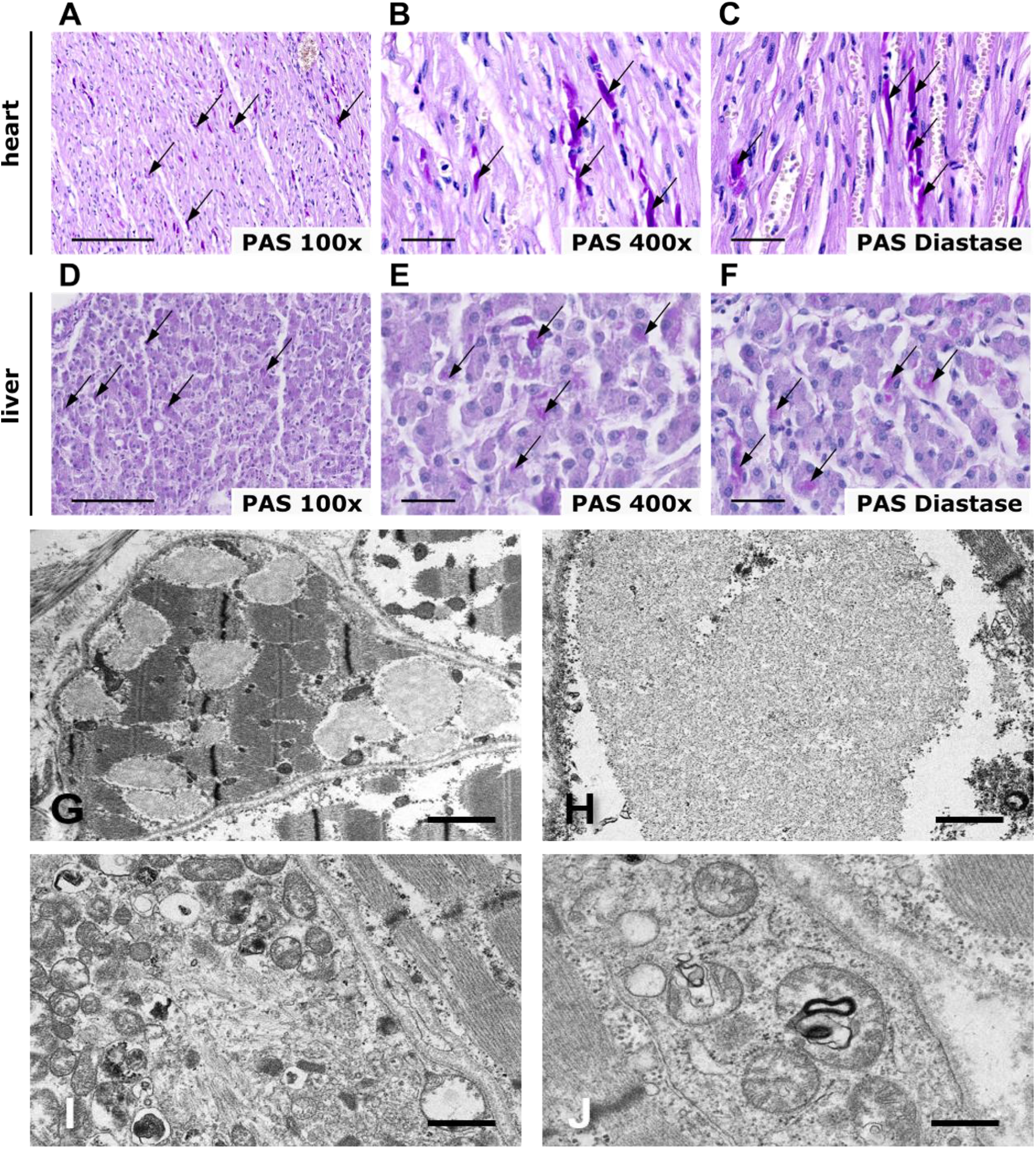
Histological findings in patient I and III. Histology of patient I revealed PAS-positive deposits in heart and liver which were still stained positive by PAS-diastase stain. (A-C) PAS-stained sections of heart muscle at 100x (A) and 400x (B, C) magnification. Liver sections at 100x (D) and 400x (E, F) magnification, before and after diastase stain. Scale bars: 200 µm (A, D), 50 µm (B, C, E, F). (G-J) Electron micrographs of the patient III muscle biopsy. (G) Muscle fiber containing several polyglucosan bodies. Scale bar = 1 µm. (H) Higher magnification of a polyglucosan body composed of finely granular and filamentous material. Scale bar = 0.4 µm. (I) Degenerating mitochondria undergoing autophagy. Scale bar = 0.5 µm. (J) Disintegrating muscle fiber mitochondria containing osmiophilic membranous material suggestive of abnormal mitophagy. Scale bar = 0.3 µm.

### C2orf69 is targeted to mitochondria by a 24 amino acid motif

To address the subcellular localization of C2orf69, the cDNA was cloned into EGFP and FLAG expression vectors. Overexpression of N-terminally EGFP-tagged C2orf69 (EGFP-C2orf69) showed a homogenous distribution in the cell (**Supplement Figure 3C**), whereas the C-terminal EGFP fusion (C2orf69-EGFP) exhibited a distinct and potentially mitochondrial localization (**Figure 3A**). The same localization was observed for FLAG-tagged (**Supplement Figure 3A-B**) and mCherry-tagged C2orf69 (**Supplement Figure 3D-E**). The mitochondrial localization of C2orf69-EGFP was confirmed by co-expression with the mitochondrial marker TOMM20-BFP. Both proteins showed robust overlap, but the C2orf69-EGFP signal was close to and not congruent with the outer mitochondrial membrane protein TOMM20, consistent with a localization to the mitochondrial matrix (**Figure 3B**). A mutant lacking the first 24 AAs of C2orf69 and a fusion protein of the SP (AA 1-24) to a C-terminal EGFP-epitope were expressed in cells. The deletion of the SP completely abolished mitochondrial localization (**Figure 3C**). However, the SP alone was sufficient to target EGFP to mitochondria (**Figure 3D**). C-terminally truncated C2orf69 isoforms mimicking the patients’ mutations (C2orf69-Δ298 (corresponding to c.298del) and C2orf69-Δ843 (corresponding to c.843_847del)) exhibited mitochondrial localization, in line with the presence of the SP being sufficient for mitochondrial targeting (**Figure 3E-F**). However, absence of C2orf69 proteins in Western blot from whole cell lysates of EBV transformed lymphocytes of patient I in contrast to control samples from unaffected family members suggested that truncated proteins are not stable, as RNA levels seemed to be not reduced (**Supplement Figure 1A, C**).

**Figure 3.**
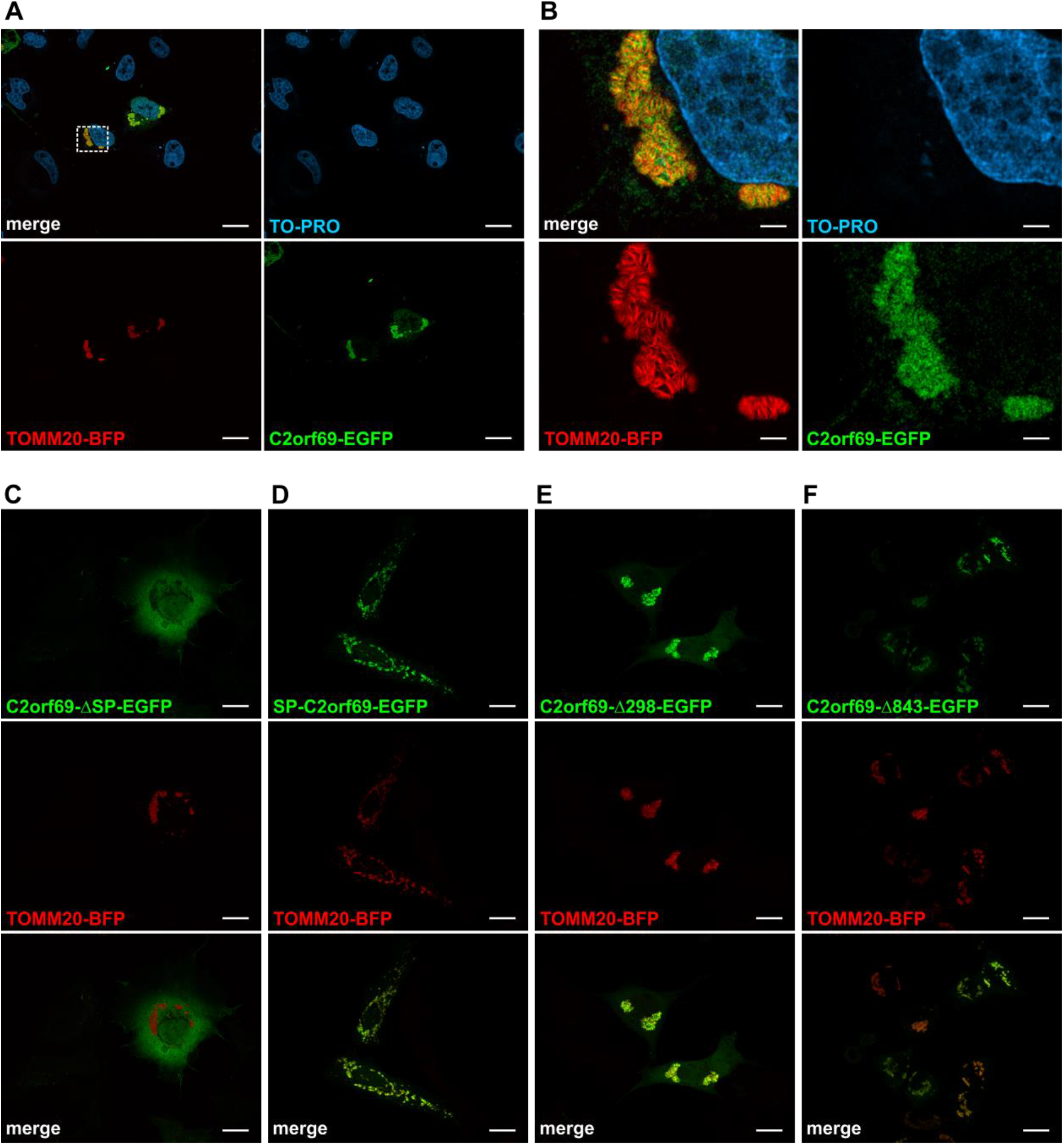
C2orf69 shows mitochondrial localization. COS-7 cells were transfected with the indicated constructs and nuclei were stained with TO-PRO. Localization of C2orf69-EGFP (A), C2orf69-ΔSP-EGFP (C), SP-C2orf69-EGFP (D), C2orf69-Δ298-EGFP (E), C2orf69-Δ843-EGFP (F), and the mitochondrial marker TOMM20-BFP (Scale bar: 20 µm). (B) Magnification of the mitochondria from A (Scale bar: 3,5 µm).

To further classify the function of *C2orf69* an NGS-based yeast-2-hybrid screen was carried out using full-length human *C2orf69* and a normalized human brain cDNA library. 35 putative interaction partners and amongst them important mitochondrial proteins (BOLA3, CMPK2, ETFA, KLHDC9, SDHB) were identified (**Supplement Figure 4**). Further, RNA-Sequencing (RNA-Seq) of EBV transformed lymphocytes of patient I and all family members revealed an upregulation of genes involved in the OXPHOS process. Moreover, an upregulation of immunological pathways like Fc receptor and TNFα signaling was observed. On the other hand, pathways related to cell proliferation e.g. histone synthesis, RNA polymerase 2 related transcription and TOR signaling were downregulated (**Supplement Figure 5**).

### C2orf69 loss causes respiratory chain defects and accumulation of reactive oxygen species

To further address the function of C2orf69, a knockout was generated by CRISPR/Cas9-based mutagenesis in HAP1 cells. The knockout (KO) in two independent C2orf69 KO clones (KO1 and KO2) was confirmed by Sanger sequencing (**Supplement Figure 6**) and Western blot (**Supplement Figure 2A-B**). Analysis of neither mitochondrial mass (**Supplement Figure 7C)** nor mitochondrial DNA copy number (**Supplement Figure 7D-F**) revealed alterations compared to WT cells. However, analyzing OXPHOS function, we detected a >20% reduction of complex I activity in both C2orf69 KO clones. Examination of OXPHOS function in a muscle biopsy of patient III also revealed a strong reduction of complex I activity (**Supplement Table 2**). In addition, a reduction of complex II activity was observed, whereas complex III and complex IV activity were not altered in the KO clones. Of note, the activity of the ATP synthase (complex V) was be impaired in C2orf69 KO cells (**Figure 4**). Another hallmark of mitochondrial malfunction is an increase in reactive oxygen species (ROS) within the cell. Therefore, cellular ROS levels were analyzed by 2,7-dichlorofluorescein (DCFDA) staining in C2orf69 WT and KO cells. DCFDA is oxidized by ROS and this results in a fluorescent form of the reagent. The analysis revealed a significantly higher amount of ROS in C2orf69 KO cells under resting conditions (**Supplement Figure 7A-B**). In summary, the data indicate OXPHOS defects which lead to alterations of complex I, II, and most robust, complex V activity.

**Figure 4.**
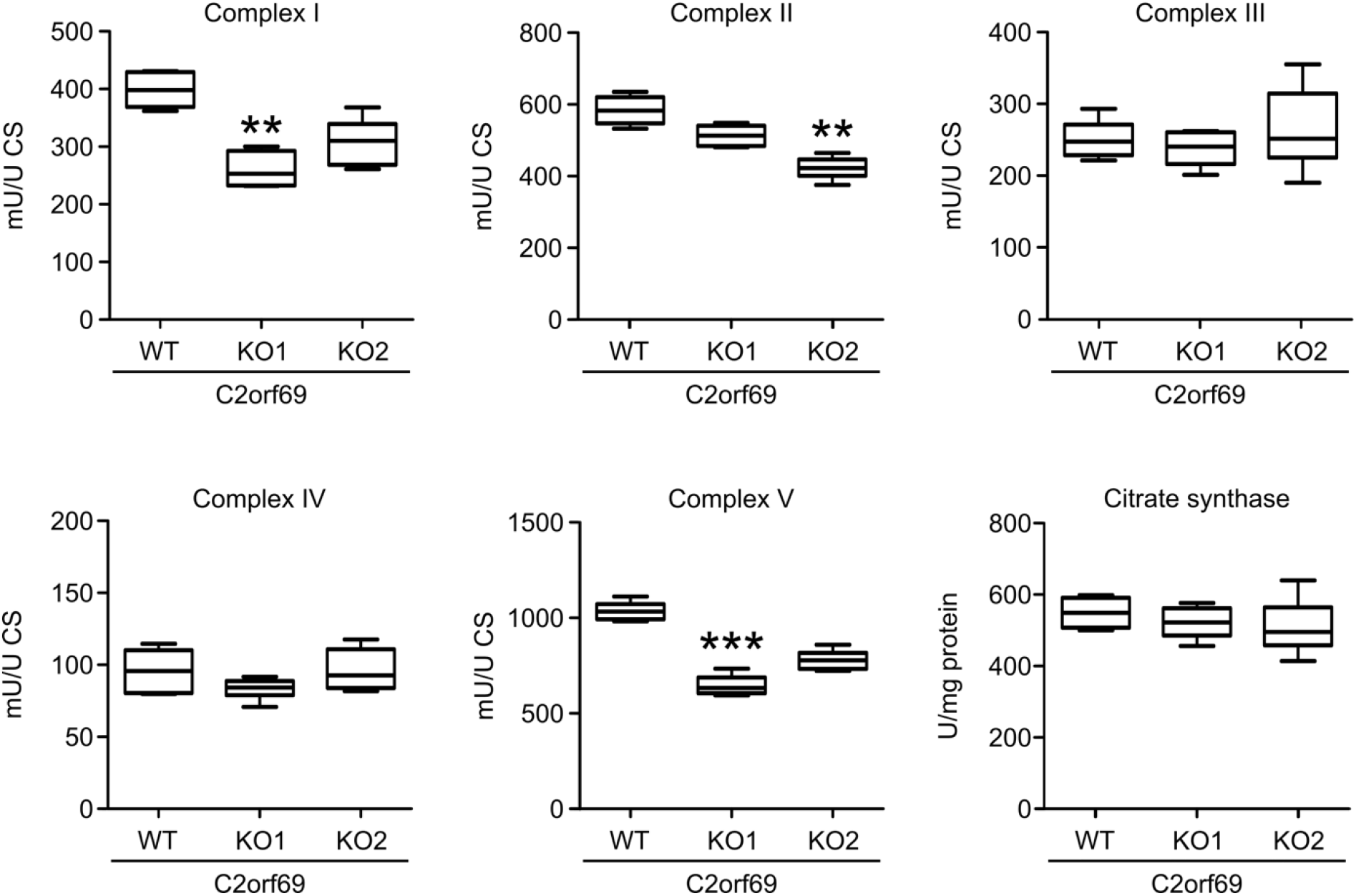
Biochemical investigations of mitochondrial respiratory chain complexes in HAP1 cells. Box plots show the mean values of WT and KO cells (n=3). Boxes show the lower and upper inter-quartile. Horizontal lines indicate the median and whiskers show min to max. Statistical significance was evaluated by Mann Whitney test with p<0.05 considered as statistically significant. **=p<0.01; ***=p<0.001, CS = citrate synthase.

## Discussion

Understanding the function of yet uncharacterized genes is a major challenge in biology. *C2orf69* is one such protein-coding gene with unknown function. It encodes a highly conserved 385 AA polypeptide without any obvious orthologs or paralogs, indicating an essential and non-redundant function. First evidence for an important biological role of *C2orf69* came from our unbiased sequencing approach in a patient with a complex disease compromising several organs and their function. By whole exome sequencing we identified a homozygous frameshift deletion (c.298del) in *C2orf69* in an affected individual which segregated with the phenotype in the family. Human genomic databases do not reveal bi-allelic *C2orf69* loss-of-function mutations, further suggesting that loss of both *C2orf69* gene copies is deleterious. The subsequent identification of homozygous loss-of-function mutations in the gene in another seven individuals with similar clinical symptoms corroborates its relevance in human disease.

We provide multiple lines of evidence that C2orf69 is a novel mitochondrial protein with impact on OXPHOS function.

Strong evidence for a mitochondrial localization is already given by *in silico* prediction tools, which on the other hand suggested that C2orf69 is secreted. Our studies showed a predominant mitochondrial localization upon overexpression of C-terminally tagged C2orf69. Co-staining with the outer mitochondrial membrane marker TOMM20 showed partial overlap and suggested that C2orf69 rather localizes to the inner membrane or mitochondrial matrix. We showed that the most N-terminal 24 AA of C2orf69 are both required and sufficient for its targeting to mitochondria and that N-terminal protein tags hinder mitochondrial targeting. N-terminal signal peptides are characteristic for mitochondrial matrix proteins and only rare or absent in proteins of other mitochondrial localization (11). Addressing the localization of endogenous C2orf69 proteins by immunohistochemistry was not possible due to a lack of suited antibodies. However, our results from heterologous studies are further strengthened by the fact that endogenous C2orf69 was found in the mitochondrial matrix by a proteomics approach (APEX MS) (12). Moreover, *in silico* predictions classify C2orf69 as oxidoreductase of the mitochondrial matrix (13). Nevertheless we cannot exclude that the protein is secreted in other cell types or under certain conditions or in addition localizes to other cell compartments than mitochondria.

Yeast-2-hybrid screening revealed 35 interaction partners of C2orf69, and several interacting proteins support its role in mitochondrial function: SDHB is a major subunit of complex II of the respiratory chain, BOLA3 is an essential protein in the Fe-S cluster synthesis and involved in the assembly of respiratory chain complexes; ETFA is involved in the electron transfers from the mitochondrial β-oxidation to the ubiquinone pool in the inner mitochondrial membrane; KLHDC9 is predicted to localize to mitochondria; CMPK2 plays an essential role in the mitochondrial nucleotide synthesis salvage pathway. Other interacting proteins could be assigned to the secretion of proteins and the extracellular matrix, indicating a possible role of C2orf69 outside the cell. However, in our secretion assays, we did not observe C2orf69 in cell supernatants. Publicly available high-throughput MS data show ubiquitination and SUMOylation of C2orf69. Consistent with this, 3 proteins involved in ubiquitination and SUMOylation are found in our Y2H screen.

There is evidence from our HAP1 C2orf69 KO model that mitochondrial complex I, II and complex V activity are reduced. The clinical data of patient III also revealed lower activity of complex I and complex IV in muscle (complex V activity was not measured). A similar impairment of complex I and IV is described in patients with Leigh syndrome which is caused by mutations in *NDUFA5* (*C20orf7*), an assembly factor of complex I (14). Immortalized lymphocytes from patient I exhibited a transcriptional upregulation of genes associated with OXPHOS compared to controls and together with increased ROS levels in the HAP1 KO cells these data corroborate mitochondrial impairment upon C2orf69 loss (15, 16). RNA-Seq data show an upregulation mainly of the mitochondrial encoded genes of complex I and III-V (**Supplement Figure 8**). This constellation is observed in response to increased ROS levels and OXPHOS alterations (17, 18). Moreover the induction of *NDUFA4* is remarkable and may be a compensatory effect, because besides being an essential part of complex I, *NDUFA4* acts as an assembly factor for complex IV (19). Further, the observed disintegration and degeneration of mitochondria in EM of patient III underlines the essential role of *C2orf69* in mitochondrial maintenance.

The clinical symptoms of the patients are in line with a multisystem mitochondrial disorder. Typical signs of mitochondriopathies include myopathy with or without cardiac involvement, deafness, various ophthalmological problems, diabetes mellitus and, in some, liver failure. Encephalopathy, seizures, cerebellar ataxia and spasticity indicate CNS affection (20). Patients with *C2orf69* mutations in accordance show muscular weakness, eye abnormalities, liver dysfunction, epilepsy and cardiomyopathy as overlapping features. Further common signs of congenital mitochondrial defects like hypomyelination/delay of myelination, cerebellar atrophy, microcephaly and short stature are also present. Lactate in blood was transiently elevated in some patients (I, III; VII) and patient III showed a lactate peak in the MR spectroscopy of the brain, however, this was not consistent for all patients.

A remarkable feature of the *C2orf69* disorder was the initial suspicion of a glycogen storage disorder (GSD) in several patients. For example, in patients III, V, VI lack of glycogen branching enzyme activity was reported and PAS-positive and diastase-resistant deposits were found in tissue biopsies (patient I, III, V, VI), features characteristic of abnormally branched glycogen as found in GSD IV. The presence of polyglucosan bodies in the EM of muscle from patient III further indicated a glycogen pathology. GSD IV is an autosomal-recessive disorder with reduced or absent GBE1 activity and a continuum of different subtypes with variable outcome ranging from fatal outcome in newborns to a late-onset neuromuscular phenotype. Early onset forms show profound hypotonia, respiratory distress, and dilated cardiomyopathy. The progressive hepatic subtype is characterized by developmental delay, a hepatomegaly, liver dysfunction, and cirrhosis accompanied by hypotonia and cardiomyopathy (21). Thus, there is marked clinical overlap between mitochondriopathies and GSD and the clinical presentation in our patients may in part be a combination of both. Inhibition of glycogen synthase (GYS1) was shown to reduce glycogen deposits in a polyglucosan body disease (APBD) mouse model (22) and thus GYS1 inhibition could be a possible option to mitigate symptoms of the *C2orf69* disorder.

A hypothetical link between mitochondrial dysfunction and glycogen accumulation could be that both pathways are involved in energy metabolism. Mitochondria are the main source of ATP, which is provided by metabolizing glucose. On the other hand, glucose is stored in the form of glycogen. The branching of the glycogen via the activity of GBE1 enables rapid synthesis and degradation to supply the cell with glucose under starving conditions. Indeed, several mitochondriopathies show signs of altered glycogen storage. The exact mechanism of the C2orf69-related pathology remains elusive, but there might be a direct or, because of the mitochondrial localization of C2orf69, rather indirect impact on GBE1 activity.

Another remarkable feature of the *C2orf69*-related disease are the recurrent inflammatory events of the patients, which were associated with an increased number of monocytes (patient I and III). In EBV-immortalized lymphocytes from patient I, several immune system related pathways seem to be disturbed. This may be in accordance with the dysregulation of the immune system in the patients, who initially seem to have an immune deficiency and later develop autoinflammation, as also seen in other mitochondrial disorders (23, 24). Therapy with glucocorticoids was partially successful in patient I, however upon reduction of glucocorticoids inflammation relapsed. Remarkably, the antiepileptic therapy with ACTH resulted in a long-lasting suppression of inflammation most probably due to the corticotropic effect. Anakinra therapy showed a remarkable improvement in patient II and the condition improved dramatically with an attenuation of serositis and arthritis and complete normalization of laboratory features. Suppression of the overacting immune system either by glucocorticoids or by targeting the interleukin-1-signalling pathway might thus be considered as therapeutic option in these patients.

The knockout of *C2orf69* in HAP1 cells revealed a reduced cell proliferation (**Supplement Figure 9**). In line with this, RNA-Seq data of family 1 also showed a downregulation of several proliferation related genes (**Supplement Figure 5**). This suggests that the attempt to compensate C2orf69 loss by upregulation of other mitochondrial genes is not sufficient and cannot provide enough energy for proliferation. Additionally, Müller *et al*. identified a CHR (cell cycle genes homology region) element in the promoter of *C2orf69*, which can be bound and repressed by the DREAM complex in response to p53 activation to regulate G2/M cell cycle genes (25, 26). C2orf69 may therefore also play a direct role in proliferation, but this has to be investigated further. Hence the impaired cell proliferation could be in part responsible for the observed clinical features, e.g. microcephaly and dystrophy.

Taken together, we here identified a novel disorder with autoinflammation as characteristic core feature and clinical overlap to mitochondriopathies and glycogen storage disorders. The affected gene product, C2orf69, localizes to mitochondria and is involved in respiratory chain function, e.g. as a complex I assembly factor, but simultaneously regulates GBE1 enzyme activity. The exact mechanism that leads to impairment of these two energy producing systems remains subject of further investigations.

## Methods

### Patients

Written informed consent was obtained from the parents / legal guardians of the study participants after approval from the Institutional Review Boards at the participating institutions (EK302-16; UK D 4886R; H12-00067, UBC). Consent was obtained according to the Declaration of Helsinki.

### Materials

If not stated otherwise, sequencing materials were ordered from Illumina, San Diego, CA, USA), cell culture reagents and cloning enzymes were ordered from Thermo Fisher Scientific (Waltham, MA, USA) and primers were ordered from biomers.net GmbH (Ulm, Germany).

### Whole-exome sequencing

Whole-exome sequencing was performed on DNA from peripheral blood of three family members including the index case and both parents (family 1). Enrichment was done with the Illumina Nextera Rapid Capture Exome v1.2 and the respective libraries were sequenced on a Illumina NextSeq500 sequencer. Alignment and variant calling were performed with SeqMule (version 1.2) (27). The resulting variant files were annotated with KGGSeq (v1.0, 14 April, 2017) (28). Variants with a minor allele frequency in public databases above 0.75 % were excluded. Mutations were confirmed by Sanger sequencing.

Exome sequencing of patient II was done as published elsewhere (29). Validation of the *C2orf69* mutation was performed by Sanger sequencing. For family 4, exome sequencing was performed from peripheral blood DNA of affected patient VI, unaffected sister and both parents. DNA Library were prepared with Illumina TruSeq DNA Library Preparation Kit. Agilent SureSelect V4 (Agilent Technologies, Santa Clara, CA, USA) was used for exome enrichment. Sequencing was performed on Illumina HiSeq 2000. Data analysis was performed using Highlander software as previously described (30): Results were confirmed by Sanger sequencing in the whole family, including patient V.

WES analysis of patient III and VI respectively VII and VIII were carried out by commercial diagnostic providers, CeGaT (Tübingen, Germany) and GeneDx (MD, USA).

### Cloning

The C2orf69 coding sequence (NM_153689.6) was amplified from a human fibroblast cDNA and cloned via *BamH*I and *EcoR*I into pEGFP-C1/N3 (Clontech Laboratories, Mountain View, CA, USA), p3xFlag-CMV-10/14 (Sigma-Aldrich, St. Louis, MO, USA) vectors. For the deletion and SP mutants the C2orf_*EcoR*I_F or C2orf69_d24_*EcoR*I-F primer and the reverse primer C2orf69_*BamH*I_del_R or C2orf69_*BamH*I_del2_R or C2orf69_*BamH*I_24AA_R was used to amplify the C2orf69 fragments from the pEGFP-C1-C2orf69 vector and cloned as for full-length C2orf69 described.

PmCherry-N1-C2orf69, pEGFP-C1-C2orf69 and pmCherry-N1 were cut by *EcoR*I and *Cfr9*I and blunted with Klenow. The resulting fragments were purified from agarose gel and ligated with T4-ligase. PmCherry-C2-C2orf69 was generated via digest of pEGFP-C1-C2orf69 and pmCherry-C2 with *BamH*I and *EcoR*I. The resulting fragments were purified via agarose gel and ligated with T4-ligase.

For the generation of px330-EGFP-C2orf69 sgRNA oligos (fw 5’ CACCGCCAGAACCATGAACCCGGG 3’ ; rev 5’ AAACCCCGGGTTCATGGTTCTGGC 3’) were cloned into the pX330-EGFP vector according to the Zhang lab protocol available from Addgene (Watertown, MA, USA). Transformation for cloning and plasmid preparation were carried out in One Shot™ Top 10 chemically competent E. *coli* (ThermoFisher Scientific, Waltham, MA, USA).

All constructed plasmids were verified by Sanger sequencing.

Mutagenesis and sequencing primers and all used vectors are listed in **Methods Table 1-3**.

**Methods Table 1.**
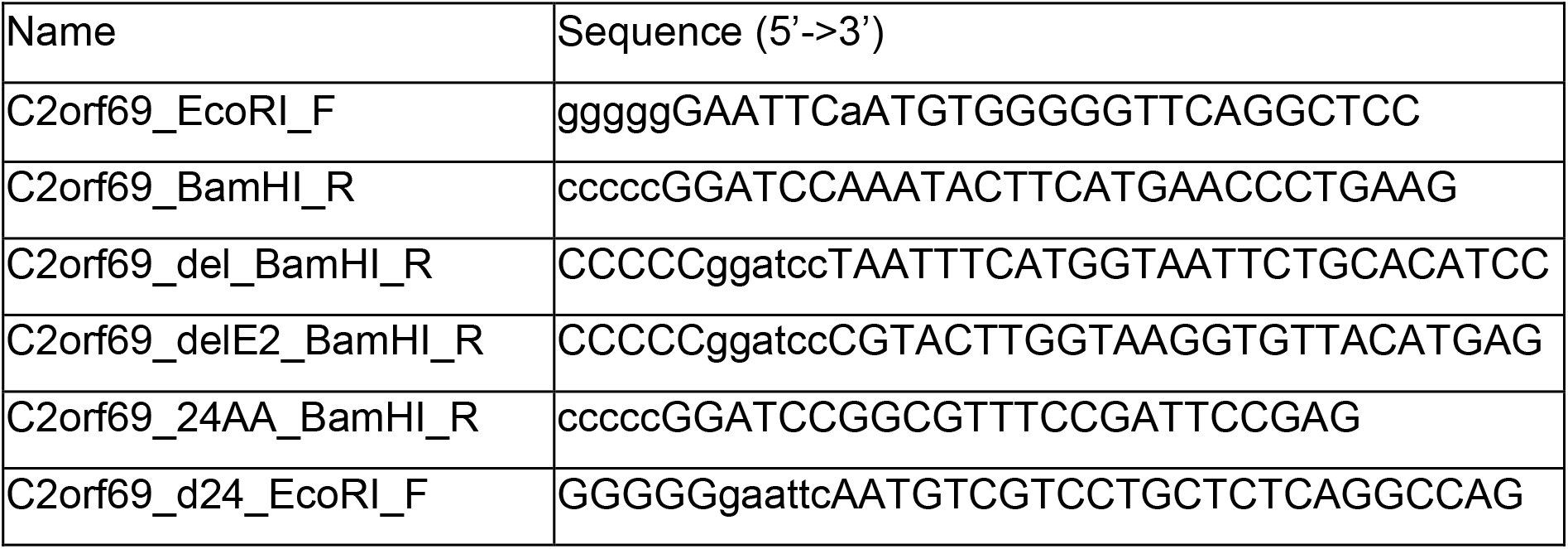
Primers for mutagenesis

**Methods Table 2.**
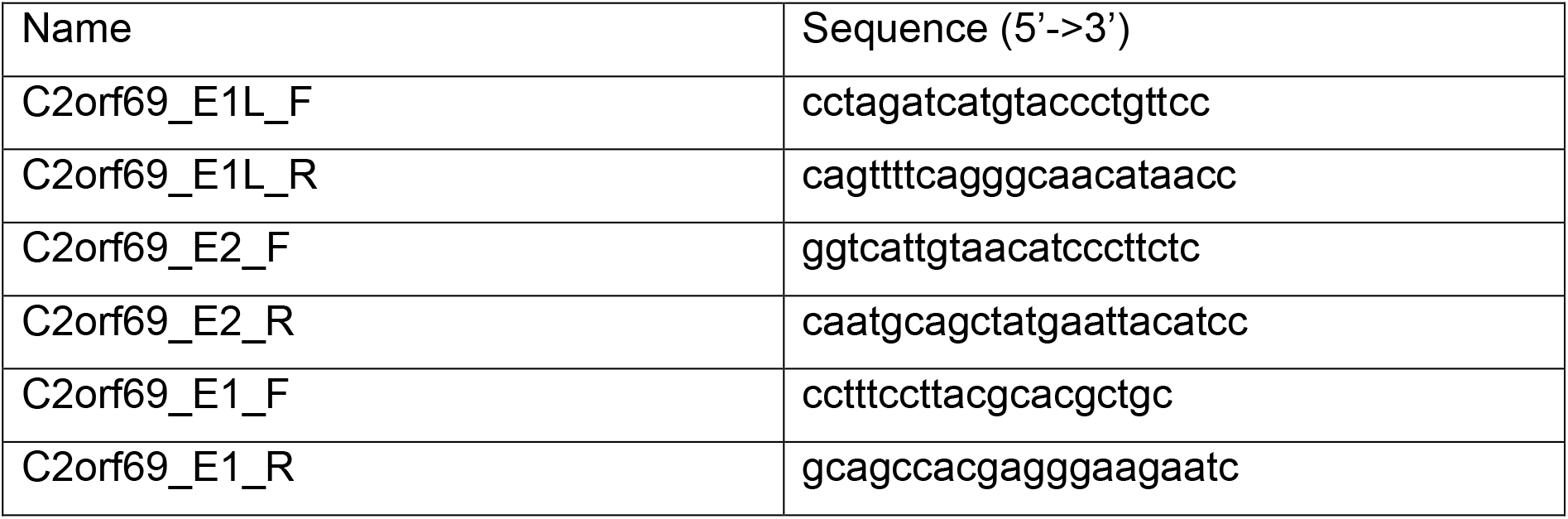
Sequencing primers

**Methods Table 3.** Vectors

C2orf69 and multisystem disorder Methods Table 3 Vectors
| Name | Backbone | Insert | Comment |
| --- | --- | --- | --- |
| pEGFP_N1 |  |  |  |
| IL6-GFP | pEGFP_N1 | IL6 | Addgene #28088 |
| pC2orf69-EGFP | pEGFP_N3 | C2orf69 | Please refer to section ***Cloning.*** |
| pEGFP-C2orf69 | pEGFP_C1 | C2orf69 | Please refer to section ***Cloning.*** |
| pC2orf69-Δ298-EGFP | pEGFP_N3 | C2orf69-Δ298 | Please refer to section ***Cloning.*** |
| pC2orf69-Δ843-EGFP | pEGFP_N3 | C2orf69-Δ843 | Please refer to section ***Cloning.*** |
| pC2orf69-ΔSP-EGFP | pEGFP_N3 | C2orf69-ΔSP | Please refer to section ***Cloning.*** |
| pC2orf69-SP-EGFP | pEGFP_N3 | C2orf69-SP | Please refer to section ***Cloning.*** |
| pC2orf69-FLAG | p3xFLAG_CMV_14 | C2orf69 | Please refer to section ***Cloning.*** |
| pFLAG-C2orf69 | p3xFLAG_CMV_10 | C2orf69 | Please refer to section ***Cloning.*** |
| pC2orf69-mCherry | pmCherry_N1 | C2orf69 | Please refer to section ***Cloning.*** |
| pmCherry-C2orf69 | pmCherry_C1 | C2orf69 | Please refer to section ***Cloning.*** |
| mTagBFP2-TOMM20-N-10 | mTagBFP2-N1 | TOMM20 | Addgene #55328 |
| pX330-U6-Chimeric_BB-CBh-hSpCas9 | pX330 |  | Addgene ID 42230 |
| pX330-U6-Chimeric-BB-C-Bh-hspCas9_EGFP | pX330-U6-Chimeric_BB-CBh-hSpCas9 | EGFP |  |
| px330-EGFP-C2orf69 | pX330 | sg RNA C2orf69 | Please refer to section ***Cloning.*** |

### Cells and cell culture

*C2orf69* knockout HAP1 cells (KO) were generated by CRISPR/Cas9. In brief, HAP1 were seeded with 70 % confluence on 6-well dishes and transfected with pX330-EGFP-C2orf69 vector. After 48 h cells were trypsinized and sorted for strong EGFP+ population. EGFP+ cells were seeded on a 10 cm dish and colonies were picked and transferred to a 96-well plate after 3 days. The knockout of the single clones was confirmed by sequencing and Western blot. The cells were cultured in Iscove’s Modified Dulbecco’s Medium (IMDM) supplemented with 5 % fetal calf serum (FCS) and 1 % penicillin/streptomycin at 37 °C in 5 % CO2 atmosphere. B lymphocytes from blood were immortalized by infection with Epstein-Barr Virus (EBV) according to Tosato *et al*. (31). HeLa (American Type Culture Collection, Manassas, VA, USA) and COS-7 (American Type Culture Collection, Manassas, VA, USA) cells were cultured in Dulbecco’s Modified Eagle’s Medium (DMEM) supplemented with 5 % FCS and 1 % penicillin/streptomycin at 37 °C in 5 % CO2 atmosphere. Transfection of HeLa, COS-7 and HAP1 cells was achieved by using PolyJet or GenJet in vitro transfection reagent (tebu-Bio GmbH, Offenbach am Main, Germany) according to the manufacturer’s protocol.

### Histology

Tissue samples, obtained during autopsy were fixed in neutral buffered formalin (4 %) followed by paraffin embedding. After sectioning, slides were stained by standard protocols with hematoxylin (Merck Millipore, Burlington, MA, USA) and eosin (Biognost, Duesseldorf, Germany), PAS (period-acid-Schiff) (Merck Millipore, Burlington, MA, USA) reaction with hematoxylin as counterstain as well as Diastase-PAS stain, similar to the PAS staining with preceding digestion with synthetic amylase (Diapath S.P.A., Martinengo BG, Italy). After coverslipping stained slides were scanned by whole-slide scanner Hamamatsu Nanozoomer 2.0HT (Hamamatsu Photonics K.K., Hamamatsu, Japan) and histological pictures were extracted by ndpview software (Hamamatsu Photonics K.K., Hamamatsu, Japan).

### Microscopy

Transfected COS-7 cells were seeded on cover slips in a 24-well plate and fixed after 24 h with 4 % paraformaldehyde in phosphate-buffered saline (PBS). TO-PRO®-3 Iodide (1:1,000; Thermo Fisher Scientific, Waltham, MA, USA) was used for nucleic acid staining. Images were taken with a Zeiss Observer Z.1 microscope (Carl Zeiss AG, Oberkochen, Germany) equipped with an Apotome2 and HXP 120 lamp.

### Yeast-two-hybrid

A commercially available Y2H screen was carried out by Next Interactions, Inc. (Richmond, CA, USA) with a Clontech human brain (normalized) library (32).

### Immunoprecipitation

HeLa cells were transfected with pEGFP-N1 (Clontech Laboratories, Mountain View, CA, USA), IL6-EGFP (Addgene, Watertown, MA, USA), pC2ORF69-EGFP or pEGFP-C2orf69 in a 6-well plate and 24 h later media was changed. After additional 6 h 500 µl of the media were collected and centrifuged, to get rid of detached cells. The supernatant was incubated with 1 µL αEGFP antibody (MAB3580, Merck Millipore, Burlington, MA, USA) for 2 h at 4 °C with agitation. 20 µL Protein A/G Bead slurry (Santa Cruz Biotechnology, Dallas, TX, USA) were added overnight, followed by three washing steps with PBS (Sigma-Aldrich, St. Louis, MO, USA) + 0.2 % Tween (AppliChem GmbH, Darmstadt, Germany). Beads were resuspended in 30 µL 2x SDS loading buffer, cooked and loaded on a SDS-gel.

### Western blot

Protein isolation and Western blot were carried out as described elsewhere (33). The primary antibodies used for immunodetection were C2orf69 (ab188870, 1:1,000, Abcam, Cambridge, UK), GFP (ab6556; 1:2,000, Abcam, Cambridge, UK), and α-tubulin (ab15246, 1:2,000, Abcam, Cambridge, UK). As secondary antibody, horseradish peroxidase-conjugated anti-rabbit (sc-2370, 1:10,000; Santa Cruz Biotechnology, Dallas, TX, USA) was used. Detection was done using Clarity Western ECL Substrate (Bio-Rad Laboratories, Inc., Hercules, CA, USA) and a FujiFilm LAS 3000 system (Fujifilm, Minato, Tokyo, Japan). Page Ruler Prestained Protein Ladder (Thermo Fisher Scientific, Waltham, MA, USA) was used for protein molecular weight estimation.

### Fluorescence-activated cell sorting

Measurements were carried out on a FACS Canto II flow cytometer (Becton Dickinson, Franklin Lakes, NJ, USA). Analysis of the data was done in FlowJo (Becton Dickinson, Franklin Lakes, NJ, USA).

### Mitochondrial Membrane Potential

Cells were seeded in 6-well plates with about 40 % confluence. 24 h later media was changed with growth media containing 200 nM Tetramethylrhodamine ethyl ester (TMRE; Biomol GmbH, Hamburg, Germany) and incubated for 20 min at 37 °C, harvested with trypsin and measured directly at FACS Canto II in the PE channel. Normalization to mitochondrial mass was carried out by parallel MitoSpy Green staining (1:10,000; BioLegend, San Diego, CA, USA).

### Reactive Oxygen Species

Cells were seeded in 6-well plates to reach about 50 % confluence at the time of the experiment. The cells were incubated with 25 µM 2’,7’-Dichlorofluorescein diacetate (DCFDA; Biomol GmbH, Hamburg, Germany) for 90 min at 37 °C, harvested with trypsin and measured directly at FACS Canto II in the FITC channel.

### Biochemical examination OXPHOS function

The activities of the OXPHOS enzyme complexes were measured in HAP cells as previously described (34, 35).

### Apoptosis

Cells were seeded in 6-well plates at 40 % confluence, harvested 24 h later with trypsin and collected in a reaction tube followed by 1 h incubation with CellEvent (2 µM in media; Thermo Fisher Scientific, Waltham, MA, USA) at 37 °C. Counterstaining was performed with 7-Aminoactinomycin D (7-AAD; 1:100; BIOZOL Diagnostica Vertrieb GmbH, Eching, Germany) which was in a solution of PBS with 5 % FCS. CellEvent and 7-AAD signal was measured by FACS in the FITC or PerCP-Cy5-5 channel, respectively.

### Growth Curve

500 HAP1 WT and KO cells were seeded on 24-well plates at day 0 in IMDM supplemented with 2 % FCS and 1 % penicillin/streptomycin at 37 °C in 5 % CO2 atmosphere. They were trypsinized and counted at day 2, 4, 6 and 8 in a Neubauer counting chamber.

### qPCR

DNA was isolated from HAP1 cells with Quick-DNA Miniprep Plus Kit (Zymo Research Europe GmbH, Freiburg, Germany) according to manufacturer’s instructions. 1 ng DNA was used for qPCR with PowerUp™ SYBR® Green (Thermo Fisher Scientific, Waltham, MA, USA) according to manufacturer’s protocol. Cycler: 2 min @95°C, 40x: 5 sec @95°C, 10 sec @60°C, 30 sec @72°C, 1 min @95°C, 1 min @60°C, ramp +0.6°C, 10 sec @95°C. Expression levels were calculated via the ΔΔCt method.

### RT-qPCR

RNA was isolated from EBV immortalized lymphocytes with peqGOLD TriFast (VWR International, Radnor, PA, USA) according to manufacturer’s instructions. 1-5 µg RNA was used for cDNA synthesis with Maxima™ First Strand cDNA Synthesis Kit (Thermo Fisher Scientific, Waltham, MA, USA) following the manufacturer’s protocol. 25 ng cDNA was used for qPCR with PowerUp™ SYBR® Green (Thermo Fisher Scientific, Waltham, MA, USA) according to manufacturer’s protocol. Cycler: 2 min @95°C, 40x: 5 sec @95°C, 10 sec @60°C, 30 sec @72°C, 1 min @95°C, 1 min @60°C, ramp +0.6°C, 10 sec @95°C. Expression levels were calculated via the ΔΔCt method.

Primers for qPCR are listed in **Methods Table 4**.

**Table 4.**
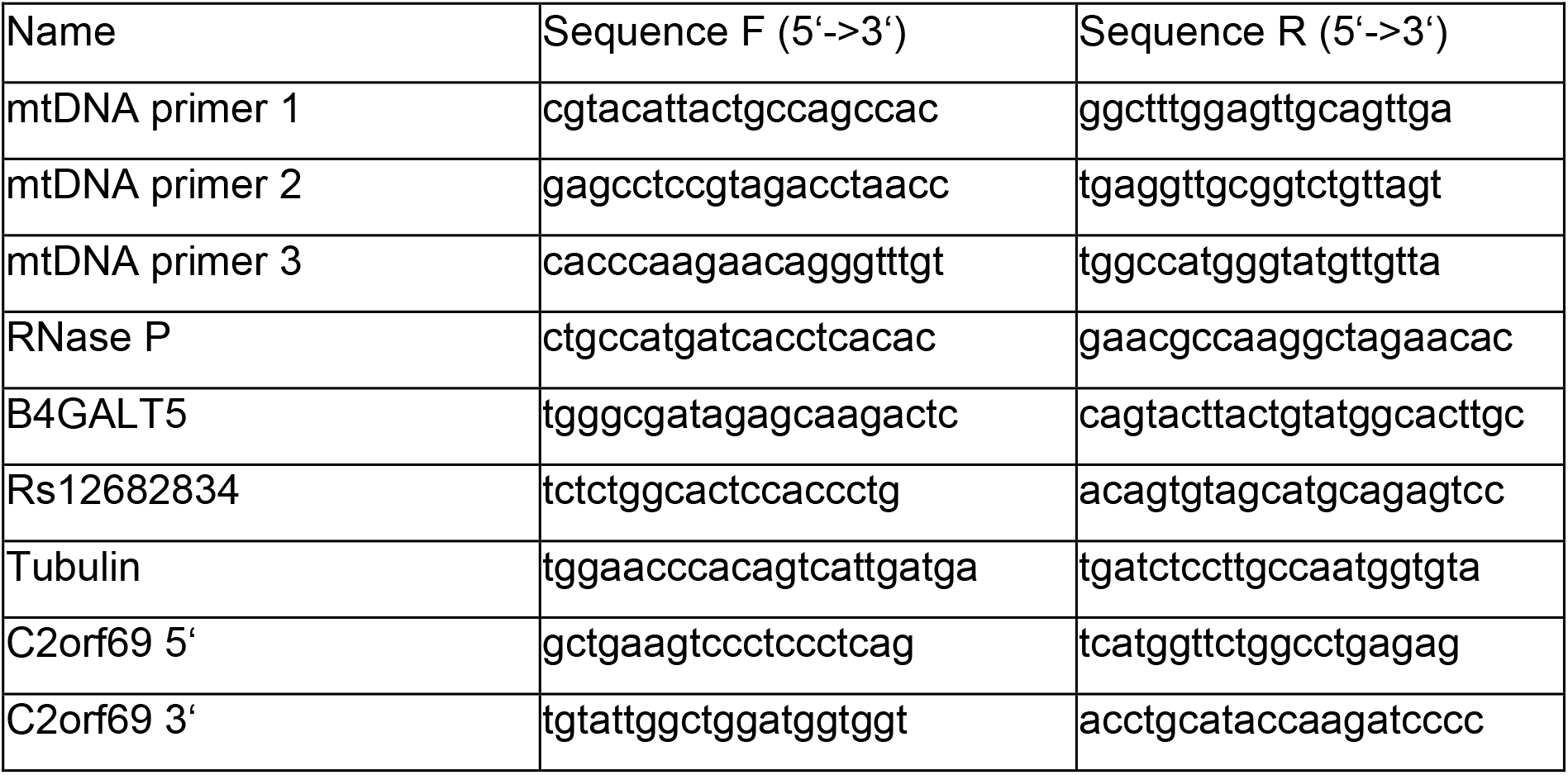
Methods qPCR primers

### Statistics

Results are presented as the mean ±SD from at least 3 independent experiments using GraphPad Prism 5. Statistical significance was evaluated by Mann Whitney test or 2way ANOVA with p<0.05 considered as statistically significant. *=p<0.05; **=p<0.01; ***=p<0.001.

### RNA-Sequencing

EBV cells were seeded with 500.000/ml 24h before RNA preparation. RNA was extracted with peqGOLD TriFast and libraries for RNA-Sequencing were prepared with the Illumina TruSeq Stranded Total RNA kit according to manufacturer’s protocol. Sequencing was carried out on a Illumina NextSeq500 with a high output 1×75 flowcell. Raw data was demultiplexed and FASTQ files were generated with bcl2fastq (v2.20). Data were analyzed and visualized with BioJupies using default parameters (36).

## Supporting information

Supplementary_Informations

## Data Availability

data is avaiable upon request, if not protected by GDPR

## Author Contributions

IK and FK design the study and analyze and interpret the data. IK, FK and EL wrote the manuscript. EL and SG acquire, analyze and interpret the majority of experimental data. PK, KO, MH, KT, ME acquire, analyze, and interpret clinical data of patient I. MB, TE, ME, RM acquire, analyzed and interpret WES data of patient I. MM analyze and interpret the MRI data of all patients. TB acquire, analyzed and interpret histology data of patient I. JW and CS acquire, analyzed and interpret EM data of patient I. PTO and AB acquire, analyze, and interpret clinical data of patient II. HS and DW acquire, analyze, and interpret WES data of patient II. CH, DC, SD acquire, analyze, and interpret clinical data of patient III. AH acquire, analyze, and interpret WES data of patient III. FGD, CL, JPD, EW and EVS acquire, analyze, and interpret clinical and WES data of patient V/VI. RS and CVK acquire, analyze, and interpret clinical and WES data of patient VII/VIII. LVDH acquire, analyze, and interpret OXPHOS data of the HAP cells. All authors carefully revise the manuscript.

## Acknowledgments

This work was funded by the Deutsche Forschungsgemeinschaft (DFG): 948/32–1 FUGG. This work was supported by the Flow Cytometry Facility, a core facility of the Interdisciplinary Center for Clinical Research (IZKF) Aachen within the Faculty of Medicine at RWTH Aachen University. Supported by a Fund Invest for Scientific Research (FIRS) Grant from the Centre Hospitalier Universitaire de Liège, Belgium. We are grateful to the TIDEX team (Dr. Britt Drogemoller, Ms. Aisha Ghani, Dr. Colin Ross, Dr. Maja Tarailo-Graovac, Dr. Wyeth Wassermann, at Centre for Molecular Medicine and Therapeutics, UBC, Vancouver, Canada). We acknowledge the physicians participating in the patients’ clinical care: Dr. Hal Siden, Canuck Place, BC Dept. of Pediatrics, Children’s Hospital, UBC, Vancouver, Canada; Dr. Michelle Demos, Dr. Lori Tucker, Department of Pediatrics, BC Children’s Hospital, UBC, Vancouver, Canada.

