## Supplementary_Informations for "A human multisystem disorder with autoinflammation, leukoencephalopathy and hepatopathy is caused by mutations in *C2orf69*"

### Supplement

### Table of contents


| | |
|---|---|
| Supplement Figure 1 | MRI in 5 patients |
| Supplement Figure 2 | Protein and RNA expression of C2orf69 in patient cells |
| Supplement Figure 3 | C2orf69 shows mitochondrial localization |
| Supplement Figure 4 | C2orf69 interacting proteins identified by NGS based Y2H screen |
| Supplement Figure 5 | RNA-Seq pathway enrichment analysis |
| Supplement Figure 6 | Sanger sequencing confirmation of C2orf69 KO cells |
| Supplement Figure 7 | C2orf69 KO alters cellular ROS levels |
| Supplement Figure 8 | RNA-Seq analysis of OXPHOS-related genes |
| Supplement Figure 9 | C2orf69 knockout reduces proliferation in HAP1 cells |
| Supplement Table 1 | Clinical description of C2orf69 patients |
| Supplement Table 2 | Biochemical findings in patient III |

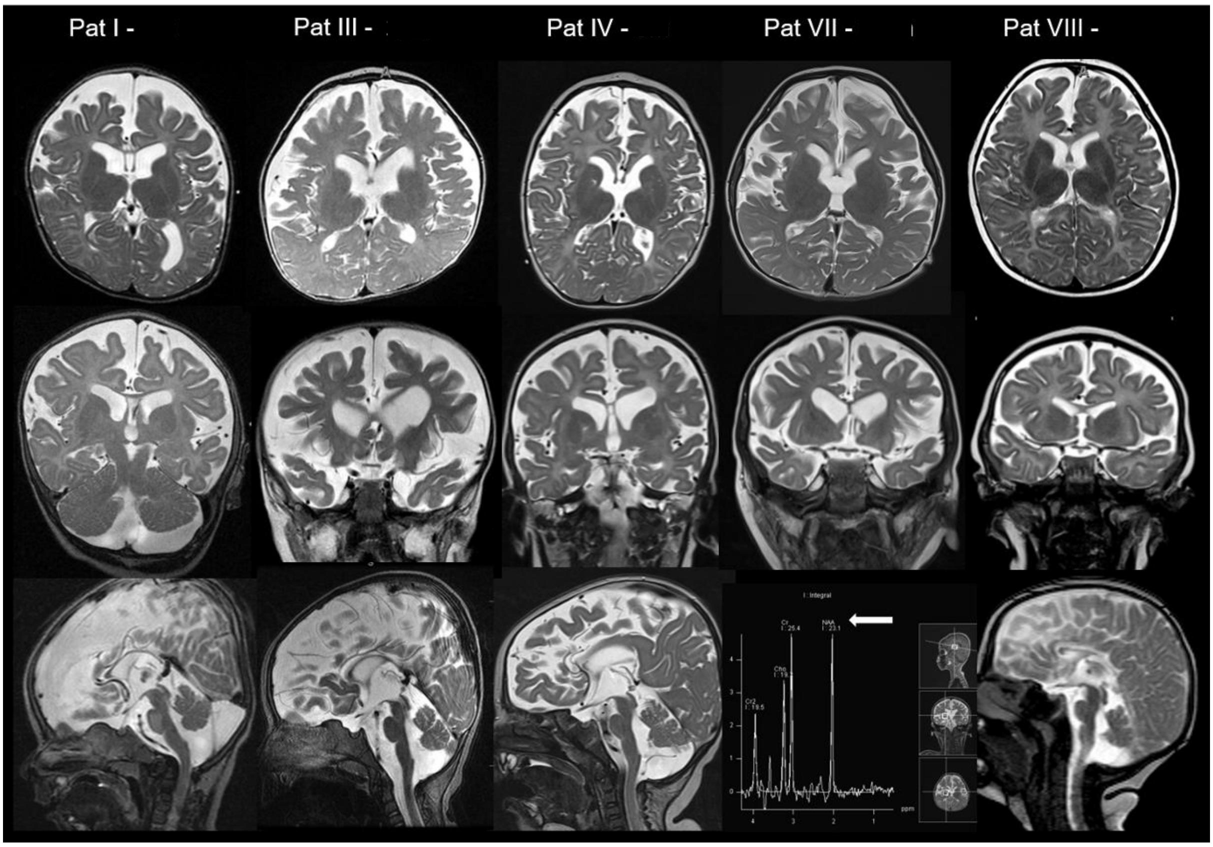


Supplement Figure 1. MRI in 5 patients. T2-weighted images in axial, coronal and sagittal orientation at different examination age. MR spectroscopy is shown in patient 7.
Different degrees of frontotemporal atrophy and hypomyelination are obvious in all patients. A thin corpus callosum and Dandy Walker variant with hypoplasia of the caudal vermis is visible in the sagittal T2-weighted images. MR spectroscopy reveals reduction of N-acetyl-aspartate in patient 7 (arrow), no prominent lactate peak can be seen.

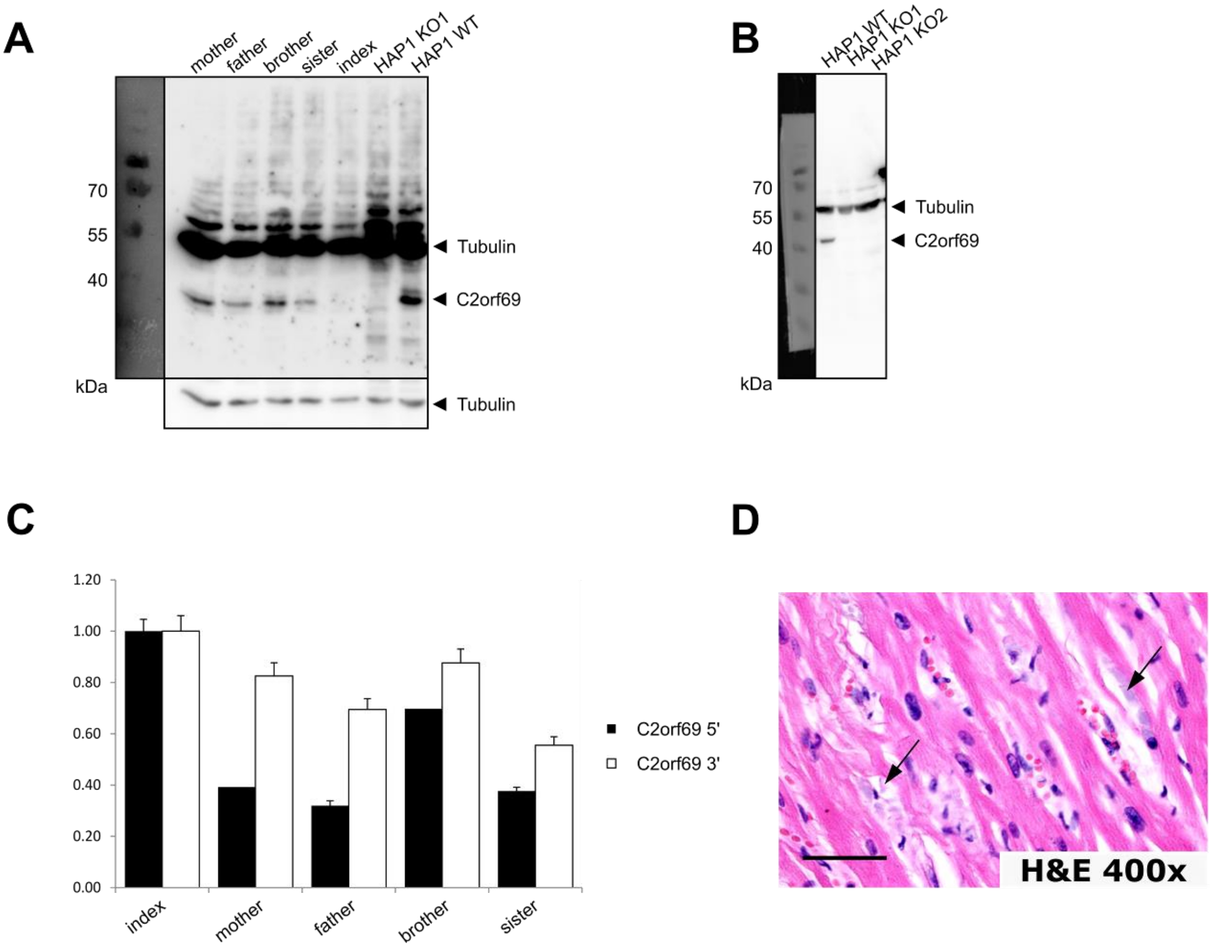


Supplement Figure 2 Protein and RNA expression of C2orf69 in patient cells. (A) Protein expression of C2orf69 in EBV cells from the index patient and his family and in HAP1 C2orf69 WT and KO cells (B) was analyzed by WB. Detection was performed with α-C2orf69 and α-tubulin-antibody. On the left side the upper panel shows the whole blot with long exposure time and corrected contrast, the lower panel shows the tubulin band with shorter exposure time. (C) qPCR of the 5' and 3' end of the C2orf69 mRNA in the index patient and his family. (D) HE staining of heart muscle in 400x magnification reveals perinuclear homogenous deposits within muscle cells. Scale bar is 50 µm.

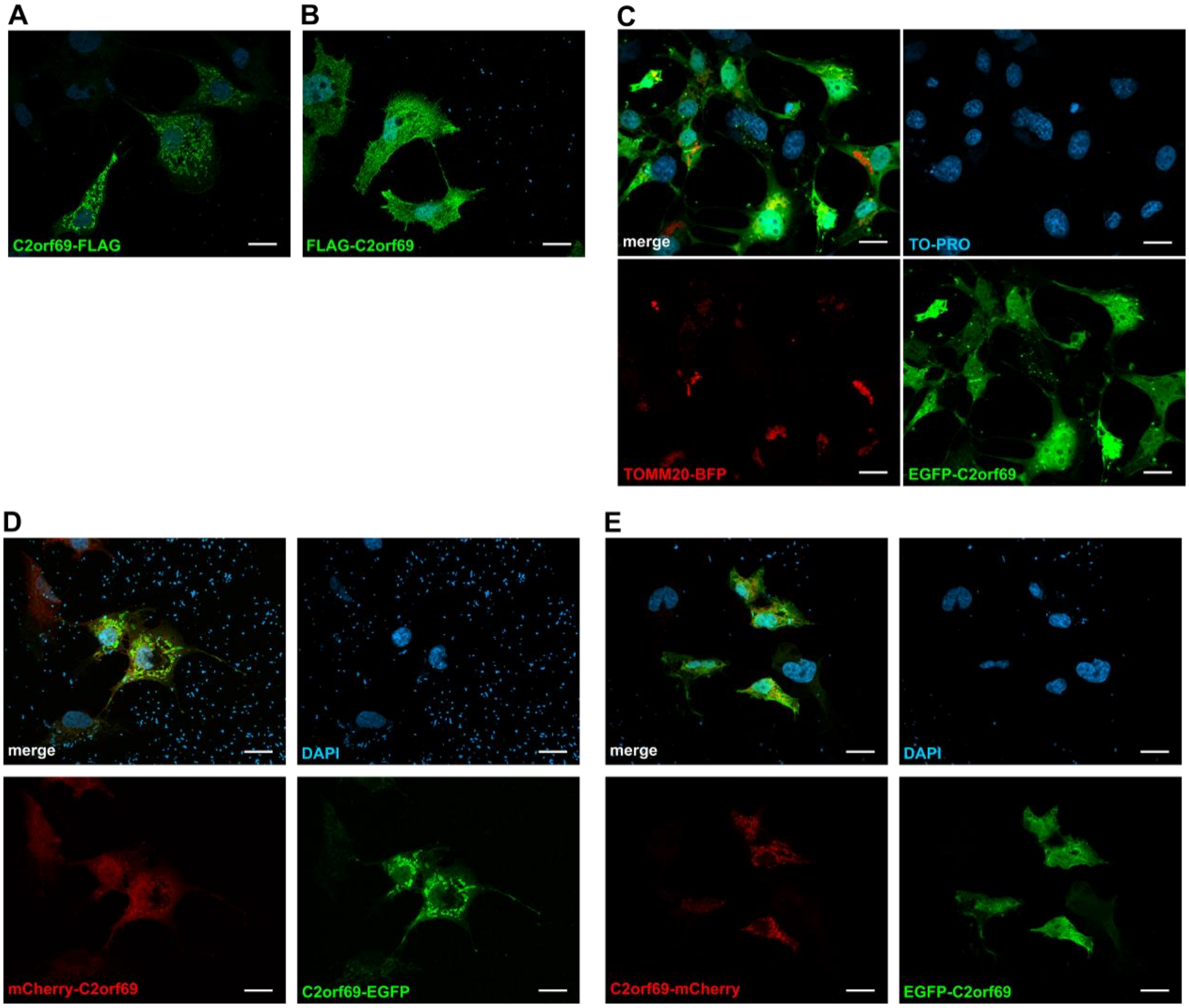


Supplement Figure 3 C2orf69 shows mitochondrial localization. COS-7 cells were transfected with the indicated constructs and nuclei were stained with DAPI (A, B, D and E) or TO-PRO (C). Localization of C2orf69-FLAG (A) or FLAG-C2orf69 (B), EGFP-C2orf69 (C), mCherry-C2orf69 and C2orf69-EGFP (D) and C2orf69-mCherry and EGFP-C2orf69 (E) (Scale bar: 20 µm).

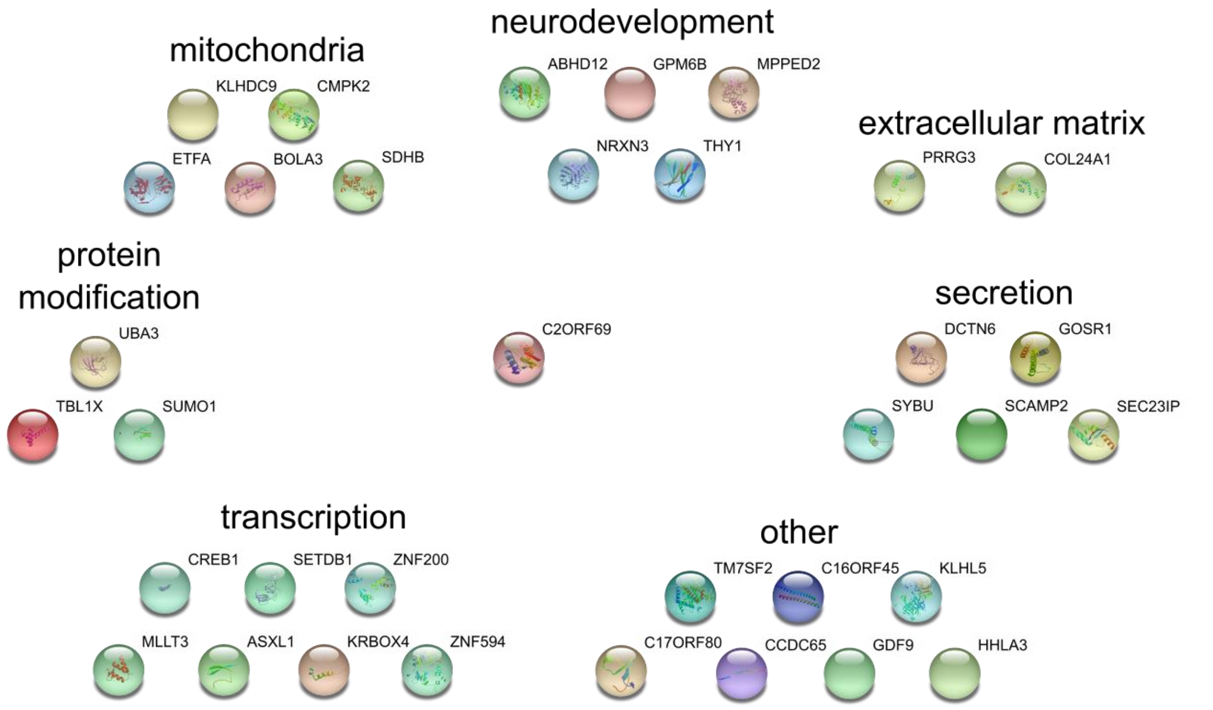


Supplement Figure 4 C2orf69 interacting proteins identified by NGS based Y2H screen and grouped by function. Modified from string.

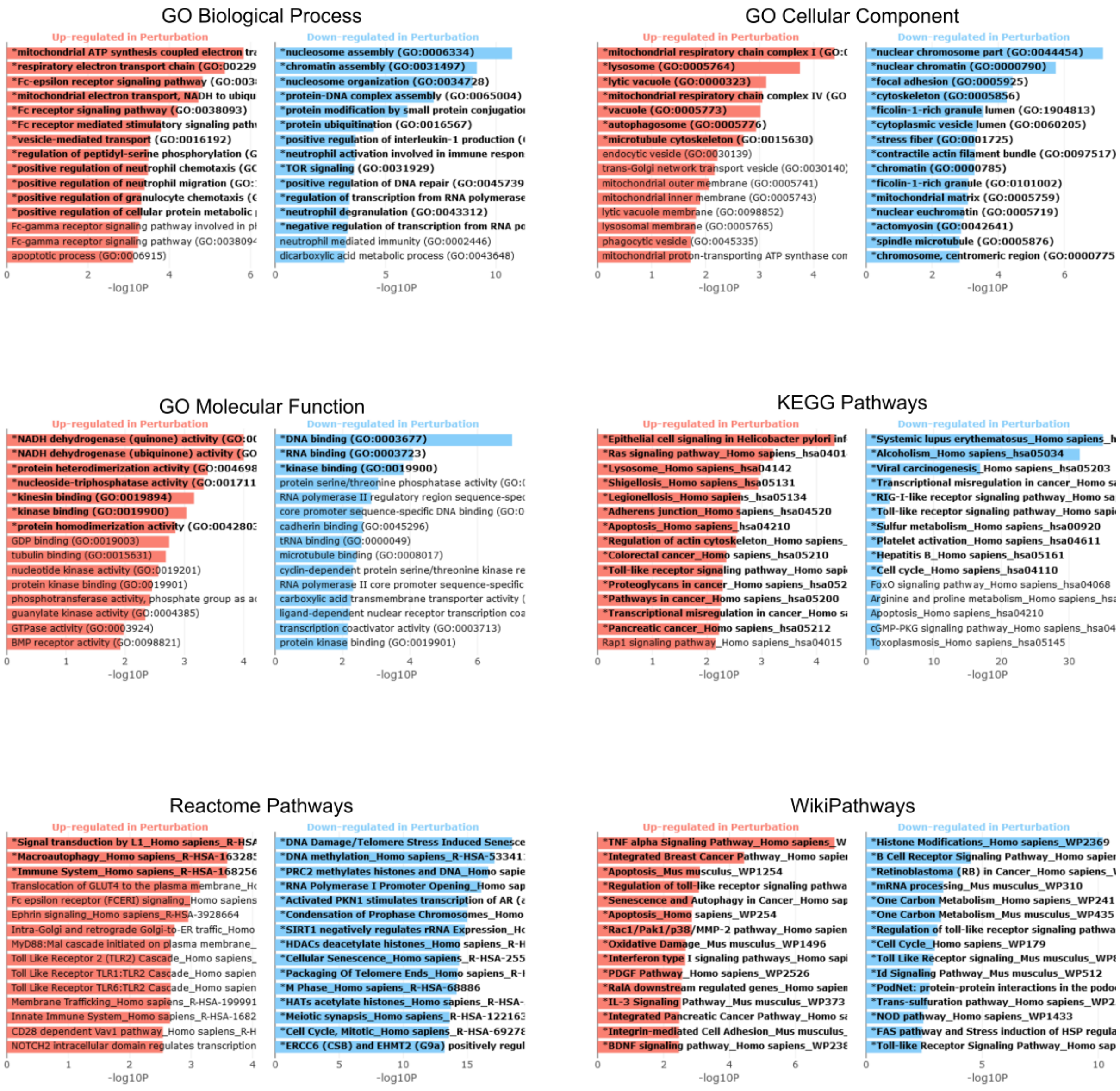


Supplement Figure 5 RNA-Seq pathway enrichment analysis. Healthy individuals of family 1 versus patient I. Significant terms are highlighted in bold.

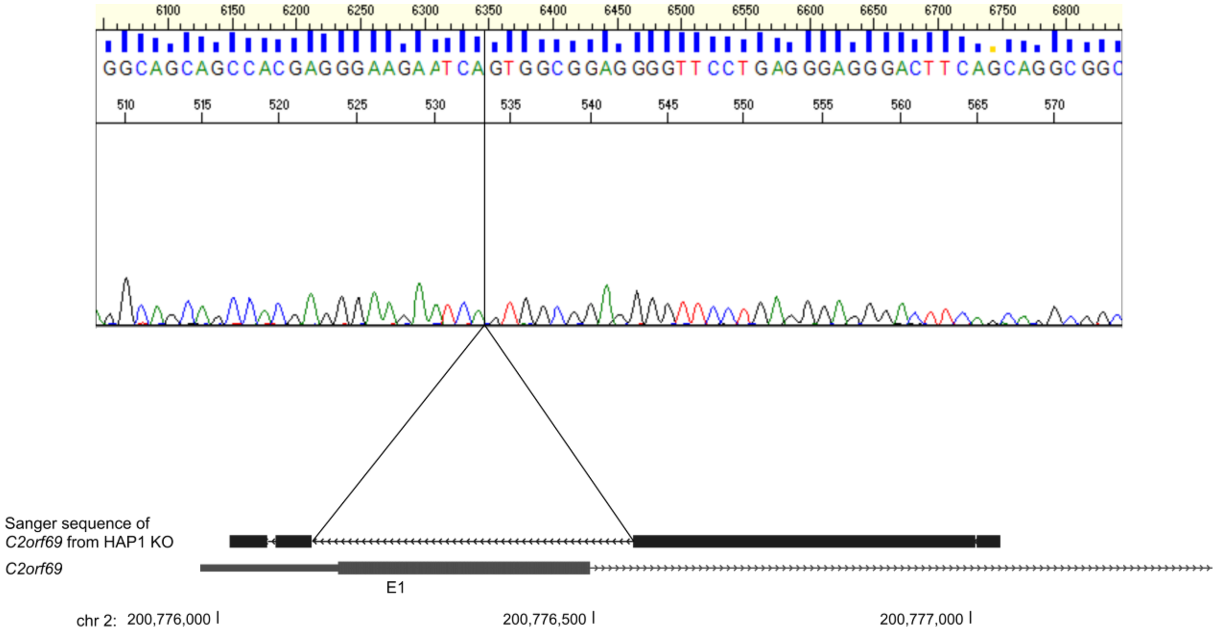


Supplement Figure 6 Sanger sequencing confirmation of C2orf69 KO cells. Sanger trace and UCSC browser view from C2orf69 sequencing analysis of HAP1 C2orf69 KO.

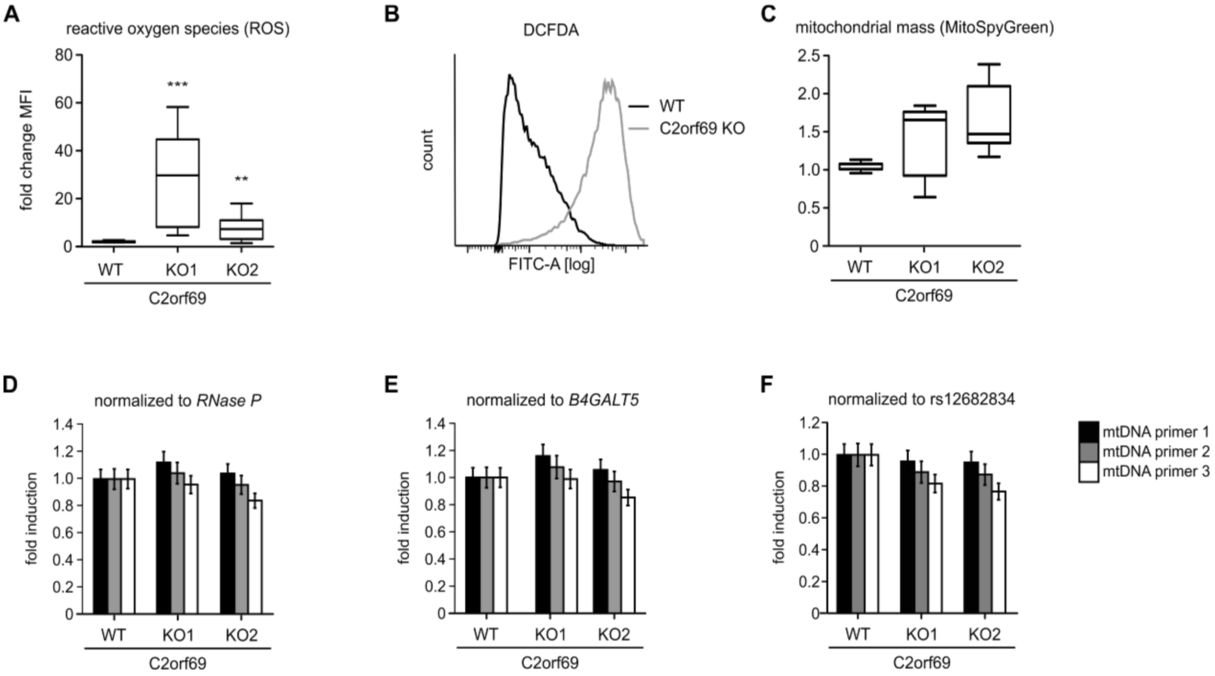


Supplement Figure 7 C2orf69 KO alters cellular ROS levels. (A) HAP1 C2orf69 WT and KO cells were stained with DCFDA and measured by flow cytometry to analyze production of ROS. Box plots show the change of mean fluorescence intensity (MFI) of WT and KO cells (n=4). Boxes show the lower and upper inter-quartile. Horizontal lines indicate the median and whiskers show min to max. All values were calculated relative to HAP1 WT cells which were set as 1. Statistical significance was evaluated by Mann Whitney test with $p<0.05$ considered as statistically significant. *=$p<0.05$; **=$p<0.01$; ***=$p<0.001$. (B) Representative curves for DCFDA. (C) MitoSpy Green staining of HAP1 C2orf69 WT and KO cells analyzed by flow cytometry to address changes in mitochondrial mass. HAP1 WT and C2orf69 KO cells were stained with MitoSpy Green and measured by flow cytometry. Quantification of the MitoSpy Green staining is illustrated by box plots (n=3). Mitochondrial plasmid amount of HAP1 WT and C2orf69 KO cells was analyzed by qPCR with three different mtDNA primers. The amount of mtDNA was normalized with quantification of genomic DNA by analysis of either RNase P (D), B4GALT5 (E) or rs12682834 (F). All values were calculated relative to HAP1 WT cells, which were set as 1. Error bars reflect SD.

**A**

**B**

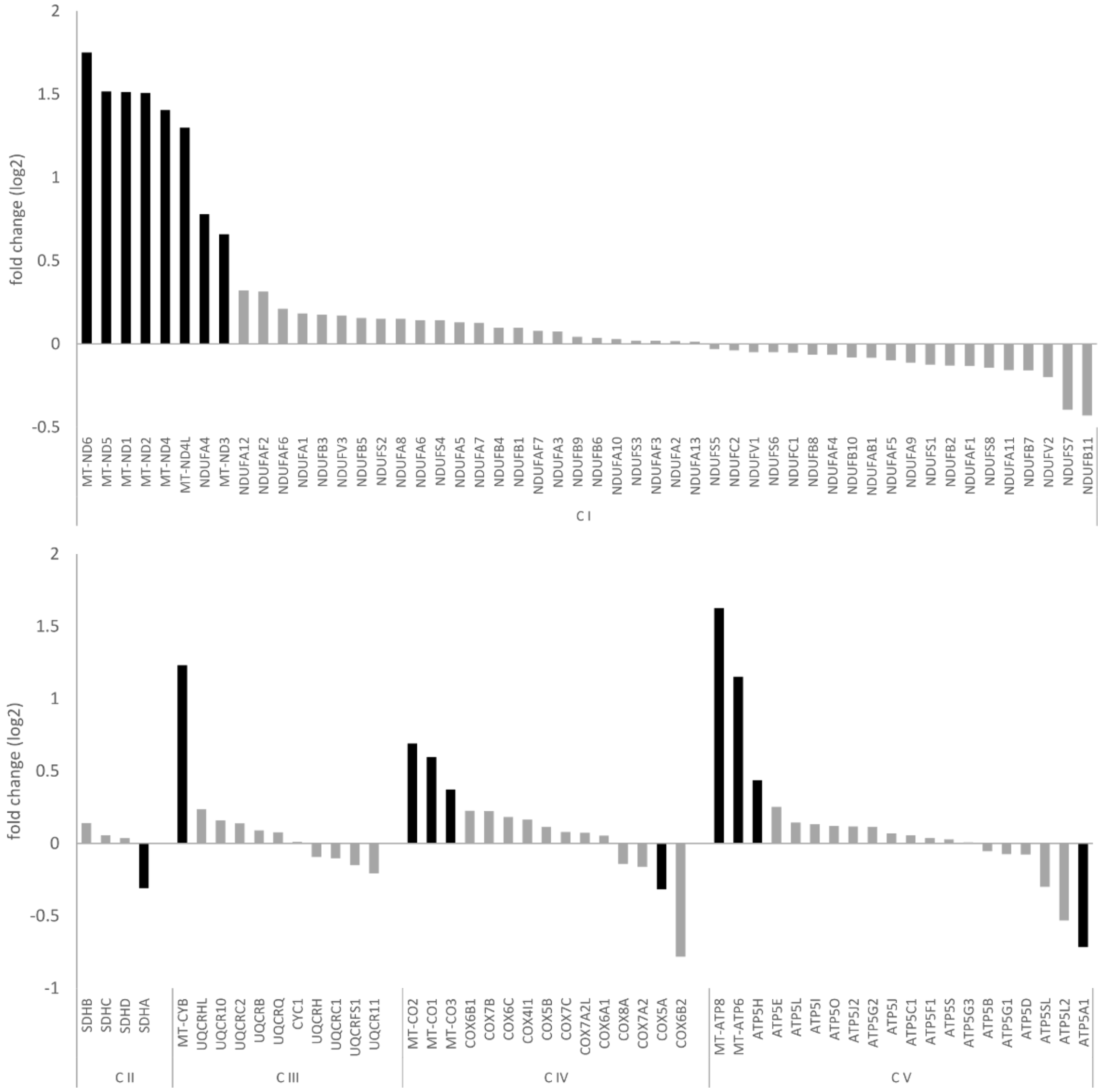


Supplement Figure 8 RNA-Seq analysis of OXPHOS-related genes. RNA-Seq results for OXPHOS and energy metabolism genes of EBV immortalized lymphocytes of patient I and his family. Black bars indicate adj. p-values <0.05, grey bars indicate not significant expression changes.

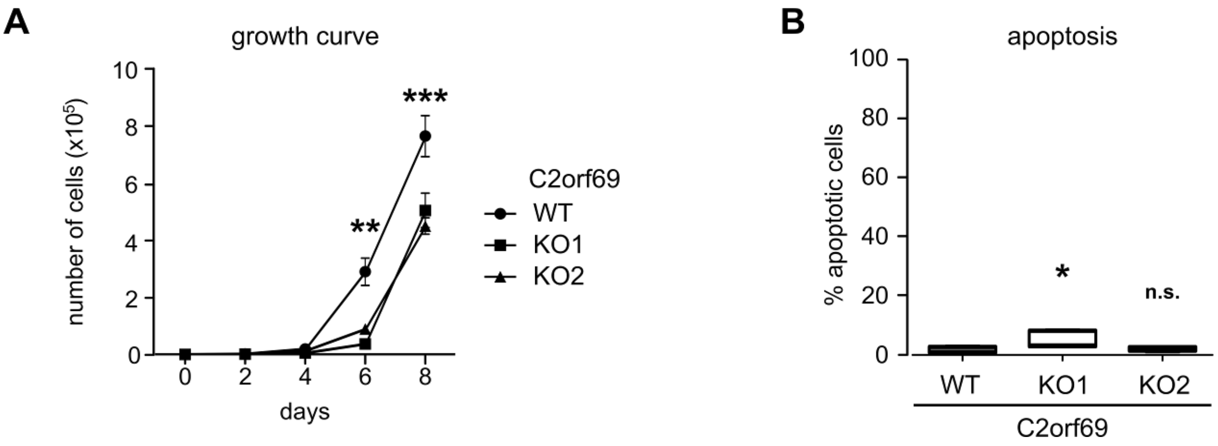


Supplement Figure 9 C2orf69 knockout reduces proliferation in HAP1 cells. (A) HAP1 WT and KO cells were seeded at day 0 and counted at the indicated time points (n=3). Error bars reflect SD. Statistical significance was evaluated by 2way ANOVA with $p<0.05$ considered as statistically significant. *=$p<0.05$; **=$p<0.01$; ***=$p<0.001$. (E) Apoptosis related caspase activity was analyzed with CellEvent in HAP1 WT and KO cells by flow cytometry (n=3) and illustrated as box plots. Boxes show the lower and upper inter-quartile. Horizontal lines indicate the median and whiskers show min to max. Statistical significance was evaluated by Mann Whitney test with $p<0.05$ considered as statistically significant. *=$p<0.05$; **=$p<0.01$; ***=$p<0.001$.

Supplement Table 1 Clinical description of C2orf69 patients

Provided as separate Excel file (clinical table.xlsx)

Supplement Table 2 Biochemical findings in patient III

| **Biochemical examination of liver biopsy** | | | | |
|---|---|---|---|---|
| | value in patient | normal value | | unit |
| | | mean | span | |
| Glycogen | 7.2 | 4.6 | 2.4-6.4 | g/100 g liver |
| Phosphorylase b-Kinase | 61.8 | 83.5 | 55.5-112.8 | U/g liver |
| 1,4-α-Glucan Branching Enzyme | 104 | 170 | ±62 | U/g liver |
| Protein (Lowry) | 800 | 181 | 141-226 | mg/g liver |
| **Biochemical examination of erythrocytes** | | | | |
| | value in patient | normal value | | unit |
| | | mean | span | |
| Phosphorylase b-Kinase | 6.7 | 4.5 | 2.1-5.5 | U/g Hb |
| Glucan Branching Enzyme | 0 | 33.2 | 18.3-50.0 | U/g Hb |
| **Biochemical examination of fibroblasts** | | | | |
| | value in patient | normal value | | unit |
| | | mean | span | |
| Beta-Glucuronidase (Reference Enzyme) | 0.49 | 1.11 | 0.47-1.83 | µmol/min/mg |
| Branching Enzyme | 0.29 | 1.32 | 1.20-1.43 | µmol/min/mg |

| **Biochemical examination of muscle biopsy** | | | |
|---|---|---|---|
| measured enzyme | value in patient | unit | normal value |
| **Respiratory chain enzyme /g NCP** | | | |
| NADH-CoQ-Oxidoreduktase (Complex I) | 52 | U/g NCP | 104-350 |
| Succinatdehydrogenase (Complex II) | - | U/g NCP | 18-48 |
| Succ. Cyt c-Oxidoreduktase (Complex II + III) | - | U/g NCP | 11-34 |
| Cytochrom c Oxidase (Complex IV) | 40 | U/g NCP | 45-121 |
| Citratsynthase (CS) | 74 | U/g NCP | 48-110 |
| **Respiratory chain enzyme / CS** | | | |
| NADH-CoQ-Oxidoreduktase (Complex I) | 1.7 | % | 6.4-21 |
| Succinatdehydrogenase (Complex II) | - | % | 22-40 |
| Succ. Cyt c-Oxidoreduktase (Complex II + III) | - | % | 15-34 |
| Cytochrom c Oxidase (Complex IV) | 54 | % | 56-115 |

NCP = non collagen protein
